# Self-perception of mental health, COVID-19 and associated sociodemographic-contextual factors in Latin American

**DOI:** 10.1101/2023.12.15.23300043

**Authors:** Pablo Roa, Guillermo Rosas, Gloria Isabel Niño Cruz, Sergio Mauricio Moreno López, Juliana Mejía Grueso, Haney Aguirre-Loaiza, Javiera Alarcón Aguilar, Rodrigo Reis, Adriano Akira Hino, Fernando López, Deborah Salvo, Andrea Ramírez Varela

**Author notes:** Correspondence: Pablo Roa.

## Abstract

**Objective:** This study aimed to estimate the prevalence of alterations in the self-perception of mental health (SpMH) during the COVID-19 pandemic and their associated factors in four Latin American countries. Methods: A cross-sectional study of data collected from adults in 2021 through the McDonnell Collaborative COVID-19 Response Survey. The sample was composed by 8125 people from Brazil, Colombia, Mexico, and Chile. A generalized linear model (GLM) for a binary outcome variable with a logistic link and fixed country effects was used. Results: There were 2336 (28.75%) people who considered having suffered alterations in SpMH. The unemployed [OR: 1.40 (95% CI: 1.24–1.58)], those with bad/regular quality of life [OR: 5.03 (95% CI: 4.01-6.31)], and those with high socioeconomic status (SES) [OR: 1.66 (95% CI: 1.41–1.96)] had a higher risk of SpMH alterations than those with full-time employment, excellent quality, and low SES status. According to the fixed-effects model, Brazilians living in the country during the pandemic, who disagreed with their government’s decisions [OR: 2.05 (95%CI: 1.74–2.42)] and lacked trust in their government [OR: 2.10 (95%CI: 1.74–2.42)] had a higher risk of having SpMH alterations. Conclusion: Nearly 30.0% of respondents indicated that the COVID-19 pandemic altered their SpMH. This outcome was associated with political, sociodemographic, and health risk factors. These findings should help policymakers develop post-pandemic community interventions.

## INTRODUCTION

The COVID-19 pandemic has had a critical impact on global public health. Governments took several measures to protect their citizens from SARS-CoV-2, such as social isolation and lockdowns, in accordance with recommendations from the World Health Organization (WHO). Even though these interventions were adopted to preserve the population, these measures disrupted people’s normal behavioral patterns and may have had an adverse effect on their mental health ^1^. Previous research showed that, under the strict lockdown, on average 10% of individuals experienced severe psychological well-being impairment, while 50% had moderate impairment ^2^. These alterations were also impacted by contextual, sociodemographic, and health factors, which influenced changes in people’s mental health following the execution of government policies, such as lockdown during the COVID-19 pandemic ^3^. Accordingly, research conducted in the context of COVID-19 revealed that mental health alterations were linked with variables such as social support, financial stability, and the availability of means to meet basic needs ^2^.

The aforementioned findings relate to people’s perceptions of the adopted measures, their trust, and their preference for governments to address the COVID-19 crisis ^4^. Considering the politicization of the public health response to COVID-19, we conjecture that voting for the incumbent and political ideology may be associated with the mental health of citizens, highlighting that political ideology corresponds to political beliefs (i.e., liberal, moderate, or conservative) and that membership in a particular political party would thus be associated with preferences for governments ^5^.

Previous research identified these variables as societal circumstances that correlate with alterations in the Self-perception of Mental Health (SpMH) ^2,4,6^, which is defined as a set of subjective beliefs about ourselves. Recognizing health experiences requires an appreciation of the individual’s subjective condition ^7^. Similarly, governments’ preventive measures to face the COVID-19 contagion are linked to their reliability, vote for the incumbent, and respondents’ political ideology. However, there is scant evidence in Latin America on how the policy decisions made by governments affected the mental health of the population, when we consider other facilitating factors. As a result, mental health was recognized as an articulated event of COVID-19 in order to inform policymakers about the identified risk factors so that they can then carry out specific interventions to protect people’s health. Consequently, this study aims to estimate the prevalence of alterations in SpMH in individuals aged 18 and older during the COVID-19 pandemic in four Latin American countries in 2020–2021, along with associated factors.

## MATERIALS AND METHODS

### Study Design and Setting

The research was designed as a cross-sectional and panel study, in which a sample was obtained by periodically surveying a population. Data were acquired in January 2021 through the McDonnell Collaborative COVID-19 Response Survey as part of the main project “Examining the influence of political ideology in mitigating COVID-19 in the Americas” ^8^. This online survey was carried out by Netquest, which built a panel of around 20.000 people in each of the four countries–Brazil, Mexico, Chile, and Colombia–under inspection. These countries were chosen because they combined Latin America’s largest countries in terms of economic growth and population size, a large COVID-19-vulnerable population and, at the time of writing, incumbent governments that ran the gamut from the populist Left (Mexico) to the populist Right (Brazil) along with governments presided over by established parties with ample governance experience ^9^. Within the contact group, online survey invitations were distributed to reach a representative sample of each country’s population by sex, age, and socioeconomic status.

### Participants and Sample Size

Non-probabilistic sampling with an automated quota system was used to collect responses similar to the sociodemographic prevalence in the four Latin American countries ^10^. We targeted individuals over 18 years who resided in one of these four Latin American countries during the 2020–2021 COVID-19 period. Observations that lacked complete information for the dependent and independent variables were eliminated. There were 169 missing values (2.03%) out of 8.125 total observations in the final sample.

### Survey description

The survey had 38 questions about government policymaking and the political ideology of respondents. COVID-19 transmission and medical care costs, post-pandemic economic growth perceptions, and attitudes toward citizens were also inquired about. There were 19 health-related and 26 social/demographic questions ^8^. All of this was implemented in a self-administered questionnaire designed to last an average of 20 to 30 minutes.

### Dependent variable

The COVID-19 pandemic has had a massive impact on public health, including mental health ^11^, which is defined as the base of emotions, reasoning, interaction, knowledge, resilience, and self-esteem. Therefore, mental health, from a public health perspective, should not be thought of in terms of a psychopathological diagnosis; it is a process that is constructed in the relationship between human beings, their emotional well-being, and their context ^12^.

In this context, the variable self-perception of mental health from the COVID-19 items about health-related behaviors was coded as a dichotomous variable (respondents can consider having alterations in their mental health or not) by means of the question: “In the past two weeks, how often do you feel negative feelings such as blue mood, despair, anxiety, depression?” (The responses to this question were divided between those who replied (often, all the time) as alterations and those who answered (hardly ever/some of the time) as non-alterations.

### Independent Variables: Socio-demographic and Health Factors

The socio-demographic and health factors considered as independent variables were: sex (decomposed into female and male categories), age groups subdivided into life stages: young adults (18 to 26), adults (27 to 59), and older adults (60 or more) ^13^, educational level (basic, intermediate, and advanced), employment status (full-time, part-time, and unemployed), physical activity (active and inactive), knowledge about COVID-19 (how confident the respondent is about knowing the ways in which COVID-19 spreads, categorized into “not sure at all”, “not very sure”, “something sure”, and “very sure”), quality of life in the pandemic (bad/regular, good, and excellent), and socioeconomic status categorized into low (1 level), medium (2 level), and high (3 level), it was categorized in this manner due to the fact that the participating nations classify their socioeconomic levels differently, making it challenging to establish standardized categories among them. Therefore, the participants were queried regarding their preferred socioeconomic status from the options provided.

### Independent variables: Contextual factors

The contextual factors considered independent variables were: trust in government, in which participants indicated whether they were neutral, trusted, or did not trust their government; effectiveness of governance strategies, in which participants indicated whether they agreed or disagreed with whether the government had implemented effective COVID-19 control strategies; political ideology, categorized as right, left, and center; and vote for the incumbent, where people indicated whether they had voted for the incumbent president. In addition, we included a four-level country factor (Mexico, Colombia, Chile, and Brazil).

Since the data was collected through a self-administered questionnaire, nine independent variables (educational level, employment status, physical activity, knowledge about COVID-19, quality of life in the pandemic, socioeconomic status, trust in government, effectiveness of governance strategies, political ideology, and vote for the incumbent) and the dependent variable SpMH were all based on the participants’ subjective impressions.

### Statistical methods

We described country and outcome characteristics using absolute and relative frequencies and proportions. The level of significance for bivariate analysis using the chi-square (χ²) independence test was 0.05 ^14^. For a multivariate statistical analysis, we employ a generalized linear model (GLM) with a dichotomous dependent variable assumed to be Bernoulli distributed and a logistic canonical link to tie its main parameter to predictors.

The factors that help to improve the Akaike information criterion (AIC) were integrated after a stepwise selection (p values less than 0.20). The odds ratio (OR) was determined using the exponent of the regression coefficient as a measure of association (see Equation 1). We used the Deviance Hypothesis Test and Wald tests to determine the final model and the individual significance of each coefficient, respectively ^15^. Furthermore, considering that country-specific contexts may alter respondents’ opinions about trust in government and approval of government interventions ^16^, we used a fixed effects model. Also, the Receiver Operating Characteristic (ROC) curve was used to evaluate the dependent variable’s sensitivity and specificity. Furthermore, we examined (based on Cook’s Distance) potentially leveraged as well as conditionally unusual data points that may be influencing the model intercept and coefficients. All variables tested had at least one significant category that may explain SpMH variations, according to the Wald test. Stepwise variable selection and AIC were also implemented.

The software R (version 4.2.1, R Foundation for Statistical Computing, Vienna, Austria) ^17^ was used to perform statistical analyses. The study obtained ethics approval from the Ethics Committee of Universidad de Los Andes, Colombia (Approval No. 202009223).

*Equation 1. Generalized linear model (GLM)*

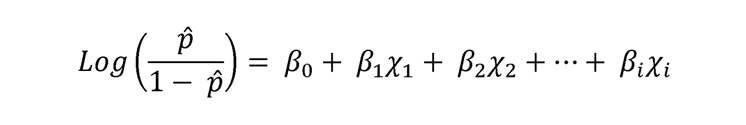

## RESULTS

### Descriptive analysis by country

Regarding socio-demographic and health characteristics, Mexico had the highest percentage of female participants (52.6%), whereas Colombia had the highest percentage of male participants (50.5%). Chile had the highest prevalence among older adults (60 and older) at 18.8%, Brazil had the highest prevalence among adults (27 to 59 years) at 72.7%, and Mexico had the highest prevalence among young adults (18 to 29 years) at 21.3%. In terms of education level, Brazil had a greater proportion of participants with basic and intermediate education (11.8% and 47.8%, respectively), whereas Colombia had a greater proportion of individuals with advanced education (80.6%). The majority of unemployed were in Chile (50.8%), while part-time workers were a plurality in Mexico (19%) and full-time workers were a plurality in Colombia (43.3%). The majority of people who assessed the quality of life during the pandemic as excellent and good were from Colombia (16.5% and 55.9%, respectively), whereas the majority of those who ranked it as bad/regular were from Chile (53.1%). According to socioeconomic status, the majority of low-level participants were from Chile (48.1%), the majority of middle-level participants were from Brazil (61.7%), and the majority of high-level participants were from Colombia (27.3%) (see Table 1).

**Table 1.**
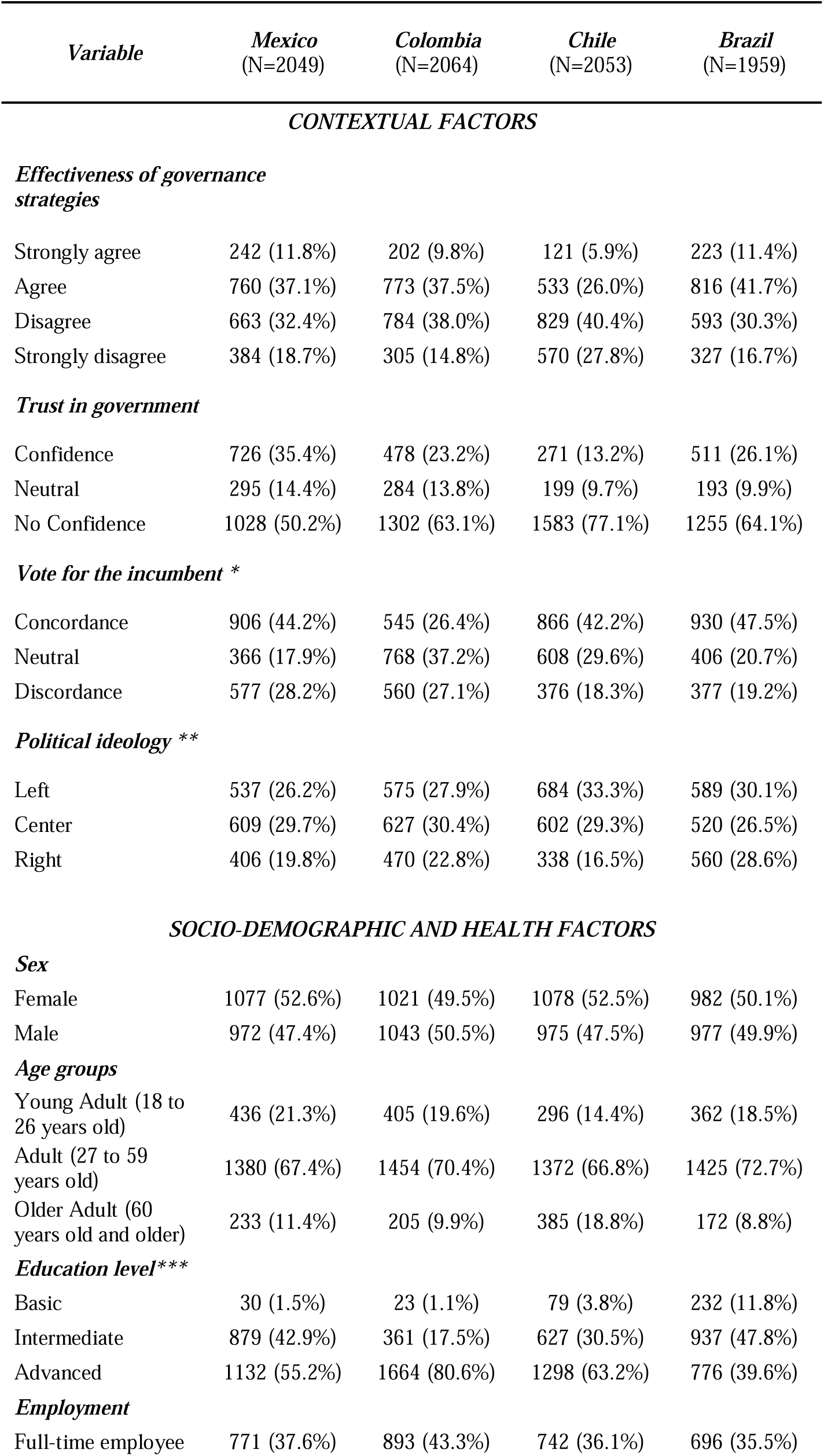
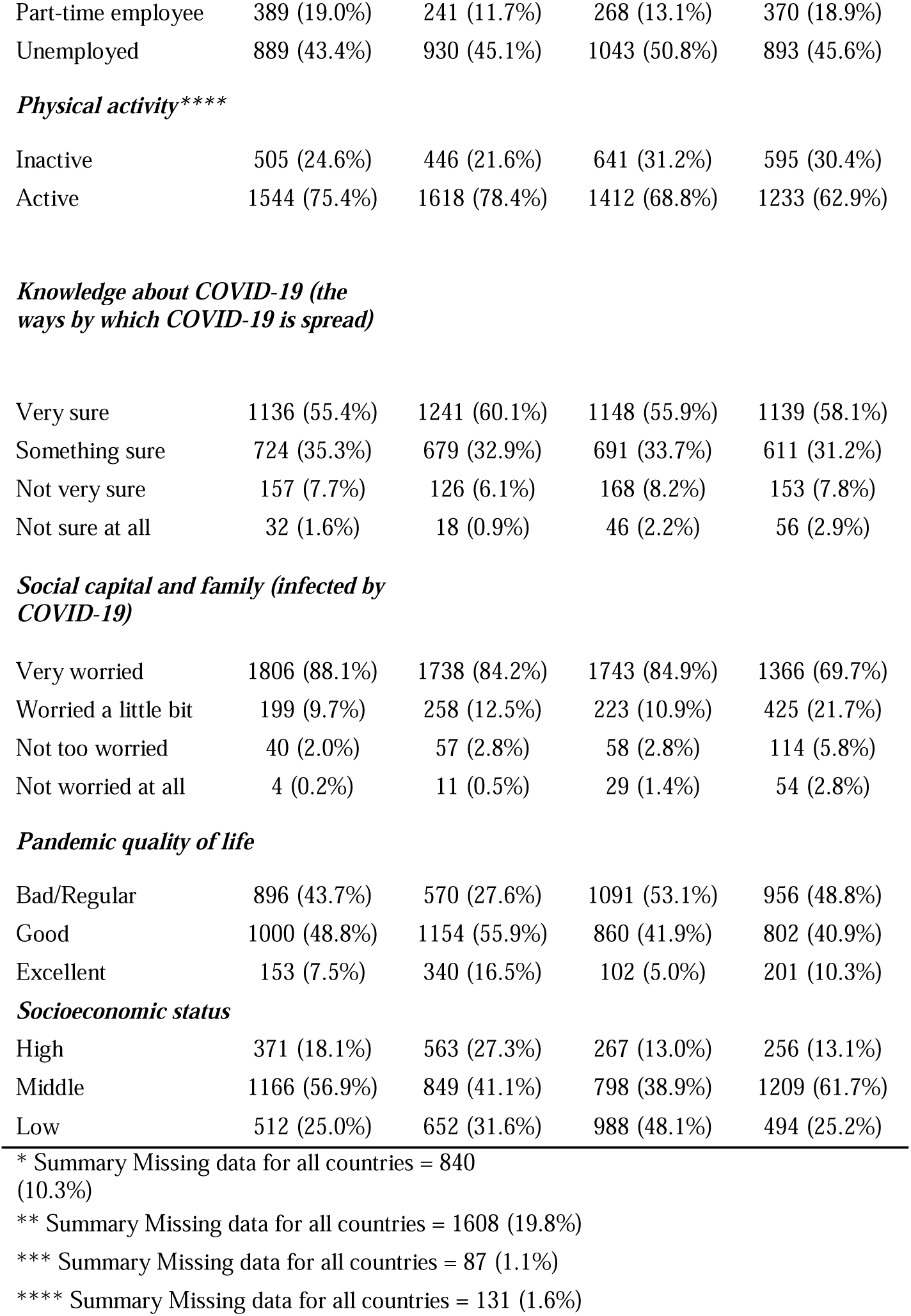
General descriptive analysis by country of Mexican, Colombian, Chilean, and Brazilian adults in January 2021.

| *Variable* | *Mexico*<br>(N=2049) | *Colombia*<br>(N=2064) | *Chile*<br>(N=2053) | *Brazil*<br>(N=1959) |
| --- | --- | --- | --- | --- |
| **CONTEXTUAL FACTORS** |  |  |  |  |
| *Effectiveness of governance strategies* |  |  |  |  |
| Strongly agree | 242 (11.8%) | 202 (9.8%) | 121 (5.9%) | 223 (11.4%) |
| Agree | 760 (37.1%) | 773 (37.5%) | 533 (26.0%) | 816 (41.7%) |
| Disagree | 663 (32.4%) | 784 (38.0%) | 829 (40.4%) | 593 (30.3%) |
| Strongly disagree | 384 (18.7%) | 305 (14.8%) | 570 (27.8%) | 327 (16.7%) |
| *Trust in government* |  |  |  |  |
| Confidence | 726 (35.4%) | 478 (23.2%) | 271 (13.2%) | 511 (26.1%) |
| Neutral | 295 (14.4%) | 284 (13.8%) | 199 (9.7%) | 193 (9.9%) |
| No Confidence | 1028 (50.2%) | 1302 (63.1%) | 1583 (77.1%) | 1255 (64.1%) |
| *Vote for the incumbent \** |  |  |  |  |
| Concordance | 906 (44.2%) | 545 (26.4%) | 866 (42.2%) | 930 (47.5%) |
| Neutral | 366 (17.9%) | 768 (37.2%) | 608 (29.6%) | 406 (20.7%) |
| Discordance | 577 (28.2%) | 560 (27.1%) | 376 (18.3%) | 377 (19.2%) |
| *Political ideology \*\** |  |  |  |  |
| Left | 537 (26.2%) | 575 (27.9%) | 684 (33.3%) | 589 (30.1%) |
| Center | 609 (29.7%) | 627 (30.4%) | 602 (29.3%) | 520 (26.5%) |
| Right | 406 (19.8%) | 470 (22.8%) | 338 (16.5%) | 560 (28.6%) |
| **SOCIO-DEMOGRAPHIC AND HEALTH FACTORS** |  |  |  |  |
| *Sex* |  |  |  |  |
| Female | 1077 (52.6%) | 1021 (49.5%) | 1078 (52.5%) | 982 (50.1%) |
| Male | 972 (47.4%) | 1043 (50.5%) | 975 (47.5%) | 977 (49.9%) |
| *Age groups* |  |  |  |  |
| Young Adult (18 to 26 years old) | 436 (21.3%) | 405 (19.6%) | 296 (14.4%) | 362 (18.5%) |
| Adult (27 to 59 years old) | 1380 (67.4%) | 1454 (70.4%) | 1372 (66.8%) | 1425 (72.7%) |
| Older Adult (60 years old and older) | 233 (11.4%) | 205 (9.9%) | 385 (18.8%) | 172 (8.8%) |
| *Education level\*\*\** |  |  |  |  |
| Basic | 30 (1.5%) | 23 (1.1%) | 79 (3.8%) | 232 (11.8%) |
| Intermediate | 879 (42.9%) | 361 (17.5%) | 627 (30.5%) | 937 (47.8%) |
| Advanced | 1132 (55.2%) | 1664 (80.6%) | 1298 (63.2%) | 776 (39.6%) |
| *Employment* |  |  |  |  |
| Full-time employee | 771 (37.6%) | 893 (43.3%) | 742 (36.1%) | 696 (35.5%) |
| Part-time employee | 389 (19.0%) | 241 (11.7%) | 268 (13.1%) | 370 (18.9%) |
| Unemployed | 889 (43.4%) | 930 (45.1%) | 1043 (50.8%) | 893 (45.6%) |
| ***Physical activity\*\*\*\**** |  |  |  |  |
| Inactive | 505 (24.6%) | 446 (21.6%) | 641 (31.2%) | 595 (30.4%) |
| Active | 1544 (75.4%) | 1618 (78.4%) | 1412 (68.8%) | 1233 (62.9%) |
| ***Knowledge about COVID-19 (the ways by which COVID-19 is spread)*** |  |  |  |  |
| Very sure | 1136 (55.4%) | 1241 (60.1%) | 1148 (55.9%) | 1139 (58.1%) |
| Something sure | 724 (35.3%) | 679 (32.9%) | 691 (33.7%) | 611 (31.2%) |
| Not very sure | 157 (7.7%) | 126 (6.1%) | 168 (8.2%) | 153 (7.8%) |
| Not sure at all | 32 (1.6%) | 18 (0.9%) | 46 (2.2%) | 56 (2.9%) |
| ***Social capital and family (infected by COVID-19)*** |  |  |  |  |
| Very worried | 1806 (88.1%) | 1738 (84.2%) | 1743 (84.9%) | 1366 (69.7%) |
| Worried a little bit | 199 (9.7%) | 258 (12.5%) | 223 (10.9%) | 425 (21.7%) |
| Not too worried | 40 (2.0%) | 57 (2.8%) | 58 (2.8%) | 114 (5.8%) |
| Not worried at all | 4 (0.2%) | 11 (0.5%) | 29 (1.4%) | 54 (2.8%) |
| ***Pandemic quality of life*** |  |  |  |  |
| Bad/Regular | 896 (43.7%) | 570 (27.6%) | 1091 (53.1%) | 956 (48.8%) |
| Good | 1000 (48.8%) | 1154 (55.9%) | 860 (41.9%) | 802 (40.9%) |
| Excellent | 153 (7.5%) | 340 (16.5%) | 102 (5.0%) | 201 (10.3%) |
| ***Socioeconomic status*** |  |  |  |  |
| High | 371 (18.1%) | 563 (27.3%) | 267 (13.0%) | 256 (13.1%) |
| Middle | 1166 (56.9%) | 849 (41.1%) | 798 (38.9%) | 1209 (61.7%) |
| Low | 512 (25.0%) | 652 (31.6%) | 988 (48.1%) | 494 (25.2%) |

\* Summary Missing data for all countries = 840 (10.3%)
\*\* Summary Missing data for all countries = 1608 (19.8%)
\*\*\* Summary Missing data for all countries = 87 (1.1%)
\*\*\*\* Summary Missing data for all countries = 131 (1.6%)

The highest prevalence of trust in government was no confidence in Chile (77.1%), followed by a neutral position and confidence in Mexico (14.4% and 35.4% respectively). In terms of vote for the incumbent, pluralities in Mexico (28.2%) and Brazil (47.5%) were discordant. Considering political ideology, self-identified rightists (28.6%) dominated in Brazil, the center (30%) in Colombia, and the left (33.3%) in Chile (see Table 1).

### Descriptive analysis by outcome

During the pandemic, 28.8% reported having alterations in SpMH and 71.2% reported not having alterations, according to the prevalence by result (see Table 2). The range of ages was from 18 to 82 years (mean ± SD: 39.9 ± 14 years). The country with the highest proportion of SpMH alterations was Brazil (32.8%), followed by Chile (26.9%), Colombia (21.0%), and Mexico (19.0%). The majority of those who said they had “no confidence” in their government (72.8%) reported the highest proportion of alterations, followed by 16.6% of those who said they had “confidence” and 10.6% of those who were neutral. According to the vote for the incumbent, “concordance” had the greatest proportion of alterations (38.4%), followed by “neutral position” (25.7%) and “discordance” (25.0%). In terms of political ideology, people who showed the highest alterations were those who identified with the left (34.8%), followed by the center (28.1%), and the right (18.1%). All the previously indicated variables were statistically significant (p<0.05 and p<0.001) (see Table 2).

**Table 2.** Descriptive analysis and χ² chi-square independence test of SpMH in Mexican, Colombian, Chilean, and Brazilian adults in January 2021.

| *Variable* | *Self-perception of mental health (SpMH)* |  | *Overall* | *P-value* |
| --- | --- | --- | --- | --- |
|  | *Without alterations in MH†* | *Alterations in MH†* |  |  |
|  | 5789 (71,24%) | 2336 (28,75%) | 8125 |  |
| **CONTEXTUAL FACTORS** |  |  |  |  |
| **Country** |  |  |  | **< 0.001** |
| Mexico | 1599 (27.6%) | 450 (19.3%) | 2049 (25.2%) |  |
| Colombia | 1573 (27.2%) | 491 (21.0%) | 2064 (25.4%) |  |
| Chile | 1425 (24.6%) | 628 (26.9%) | 2053 (25.3%) |  |
| Brazil | 1192 (20.6%) | 767 (32.8%) | 1959 (24.1%) |  |
| **Effectiveness of governance strategies** |  |  |  | **< 0.001** |
| Strongly agree | 607 (77.0%) | 181 (23.0%) | 788 (9.7%) |  |
| Agree | 2173 (75.4%) | 709 (24.6%) | 2882 (35.5%) |  |
| Disagree | 1980 (69.0%) | 889 (31.0%) | 2869 (35.3%) |  |
| Strongly disagree | 1029 (64.9%) | 557 (35.1%) | 1586 (19.5%) |  |
| **Trust in government** |  |  |  | **< 0.001** |
| Confidence | 1598 (27.6%) | 388 (16.6%) | 1986 (24.4%) |  |
| Neutral | 724 (12.5%) | 247 (10.6%) | 971 (12.0%) |  |
| No Confidence | 3467 (59.9%) | 1701 (72.8%) | 5168 (63.6%) |  |
| **Vote for the incumbent \*** |  |  |  | **< 0.05** |
| Concordance | 2350 (40.6%) | 897 (38.4%) | 3247 (40.0%) |  |
| Neutral | 1544 (26.7%) | 604 (25.9%) | 2148 (26.4%) |  |
| Discordance | 1305 (22.5%) | 585 (25.0%) | 1890 (23.3%) |  |
| **Political ideology \*\*** |  |  |  | **< 0.001** |
| Left | 1573 (27.2%) | 812 (34.8%) | 2385 (29.4%) |  |
| Center | 1701 (29.4%) | 657 (28.1%) | 2358 (29.0%) |  |
| Right | 1343 (23.2%) | 431 (18.5%) | 1774 (21.8%) |  |
| **SOCIO-DEMOGRAPHIC AND HEALTH FACTORS** |  |  |  |  |
| **Sex** |  |  |  | **< 0.001** |
| Female | 2766 (47.8%) | 1392 (59.6%) | 4158 (51.2%) |  |
| Male | 3023 (52.2%) | 944 (40.4%) | 3967 (48.8%) |  |
| **Age groups** |  |  |  | **< 0.001** |
| Young Adult (18 to 26 years old) | 860 (14.9%) | 639 (27.4%) | 1499 (18.4%) |  |
| Adult (27 to 59 years old) | 4107 (70.9%) | 1524 (65.2%) | 5631 (69.3%) |  |
| Older Adult (60 years old and older) | 822 (14.2%) | 173 (7.4%) | 995 (12.2%) |  |
| **Education level\*\*\*** |  |  |  | **0.209** |
| Basic | 255 (4.4%) | 109 (4.7%) | 364 (4.5%) |  |
| Intermediate | 2018 (34.9%) | 786 (33.6%) | 2804 (34.5%) |  |
| Advanced | 3451 (59.6%) | 1419 (60.7%) | 4870 (59.9%) |  |
| ***Employment*** |  |  |  | **< 0.001** |
| Full-time employee | 2335 (40.3%) | 767 (32.8%) | 3102 (38.2%) |  |
| Part-time employee | 926 (16.0%) | 342 (14.6%) | 1268 (15.6%) |  |
| Unemployed | 2528 (43.7%) | 1227 (52.5%) | 3755 (46.2%) |  |
| ***Physical activity\*\*\*\**** |  |  |  | **< 0.001** |
| Inactive | 1474 (25.5%) | 713 (30.5%) | 2187 (26.9%) |  |
| Active | 4215 (72.8%) | 1592 (68.2%) | 5807 (71.5%) |  |
| ***Knowledge about COVID-19 (the ways by which COVID-19 is spread)*** |  |  |  | 0.700 |
| Very sure | 3362 (58.1%) | 1302 (55.7%) | 4664 (57.4%) |  |
| Something sure | 1914 (33.1%) | 791 (33.9%) | 2705 (33.3%) |  |
| Not very sure | 410 (7.1%) | 194 (8.3%) | 604 (7.4%) |  |
| Not sure at all | 103 (1.8%) | 49 (2.1%) | 152 (1.9%) |  |
| ***Social capital and family (infected by covid 19)*** |  |  |  | **< 0.01** |
| Very worried | 4676 (80.8%) | 1977 (84.6%) | 6653 (81.9%) |  |
| Worried a little bit | 843 (14.6%) | 262 (11.2%) | 1105 (13.6%) |  |
| Not too worried | 191 (3.3%) | 78 (3.3%) | 269 (3.3%) |  |
| Not worried at all | 79 (1.4%) | 19 (0.8%) | 98 (1.2%) |  |
| ***Pandemic quality of life*** |  |  |  | **< 0.001** |
| Bad/Regular | 2017 (34.8%) | 1496 (64.0%) | 3513 (43.2%) |  |
| Good | 3064 (52.9%) | 752 (32.2%) | 3816 (47.0%) |  |
| Excellent | 708 (12.2%) | 88 (3.8%) | 796 (9.8%) |  |
| ***Socioeconomic status*** |  |  |  | **< 0.001** |
| High | 1033 (17.8%) | 424 (18.2%) | 1457 (17.9%) |  |
| Middle | 2793 (48.2%) | 1229 (52.6%) | 4022 (49.5%) |  |
| Low | 1963 (33.9%) | 683 (29.2%) | 2646 (32.6%) |  |

\* Summary Missing data for all countries = 840 (10.3%)
\*\* Summary Missing data for all countries = 1608 (19.8%)
\*\*\* Summary Missing data for all countries = 87 (1.1%)
\*\*\*\* Summary Missing data for all countries = 131 (1.6%)
MH†: Mental Health

Regarding socio-demographic and health characteristics, the unemployed related the greatest alterations in SpMH (52.5%), followed by full-time employees (32.8%) and part-time employees (14.6%). The majority of people with alterations in SpMH ranked the pandemic quality of life index as bad/regular (64%) followed by those who rated it as good (32.2%) and excellent (3.8%). Participants with a middle socioeconomic status had the highest prevalence of alterations (52.6%), followed by those with a low socioeconomic status (29.2%) and those with a high socioeconomic status (18.2%).

The variables education level and knowledge about COVID-19 were not significant (p>0.05). The relationships between the other variables and the outcome were statistically significant (p<0.001 and p<0.01) (see table 2).

### Factors associated with alterations in SpMH

People who lived in Brazil during the COVID-19 pandemic [OR: 2.5 (95%CI: 2.18–3.0)] were more likely to have alterations in the SpMH than those who lived in other countries in the region, such as Mexico. People who self-identified as “no confidence” [OR: 1.15 (95%CI: 0.97-1.37)], “discordance” [OR: 1.21 (95%CI: 1.04-1.41)], and “left-wing” [OR: 1.20 (95%IC: 1.05-1.38)] had more chances to have alterations in the SpMH than neutral and right-wing individuals (see Table 3).

**Table 3.** Inferential Analysis, General Linear Model, and Wald Test of SpMH with 5.459 total observations excluding missing data in Mexican, Colombian, Chilean, and Brazilian adults in January 2021.

| *Variables* | *Model 1 (Theoretical)* |  |  |  |  |  | *Model 2 (Saturated)* |  |  |
| --- | --- | --- | --- | --- | --- | --- | --- | --- | --- |
|  | *OR Unadj* | *CI 95%* | *Pr(> \|z\|)* | *OR adj* | *CI 95%* | *Pr(> \|z\|)* | *OR adj* | *CI 95%* | *Pr(> \|z\|)* |
| **CONTEXTUAL FACTORS** |  |  |  |  |  |  |  |  |  |
| *Country* |  |  |  |  |  |  |  |  |  |
| Mexico | 1 |  |  | 1 |  |  | 1 |  |  |
| Colombia | 1.1 | 0.9-1.2 | < **0.05** | 1.3 | 1.1-1.5 | < **0.001** | 1.3 | 1.1-1.6 | < **0.001** |
| Chile | 1.6 | 1.3-1.8 | < **0.001** | 1.4 | 1.2-1.7 | < **0.001** | 1.5 | 1.3-1.8 | < **0.001** |
| Brazil | 2.2 | 1.9-2.6 | < **0.001** | 2.5 | 2.1-2.9 | < **0.001** | 2.5 | 2.1-3.0 | < **0.001** |
| *Effectiveness of governance strategies* |  |  |  |  |  |  |  |  |  |
| Strongly agree | 1 |  |  | 1 |  |  | 1 |  |  |
| Agree | 1.09 | 0.9-1.3 | 0.2 | 0.8 | 0.7-1.09 | 0.2 | 0.8 | 0.7-1.09 | 0.2 |
| Disagree | 1.5 | 1.2-1.8 | 0.3 | 1.1 | 0.8-1.3 | 0.3 | 1.1 | 0.8-1.3 | 0.3 |
| Strongly disagree | 1.8 | 1.4-2.2 | < **0.05** | 1.2 | 1.03-1.6 | < **0.05** | 1.2 | 1.02-1.6 | < **0.05** |
| *Trust in government* |  |  |  |  |  |  |  |  |  |
| Neutral | 1 |  |  | 1 |  |  | 1 |  |  |
| Confidence | 0.7 | 0.5-0.8 | < **0.05** | 0.8 | 0.6-1.0 | 0.1 | 0.8 | 0.6-0.9 | 0.1 |
| No Confidence | 1.4 | 1.2-1.6 | < **0.001** | 1.1 | 0.9-1.3 | < **0.05** | 1.1 | 0.9-1.3 | < **0.05** |
| *Vote for the incumbent* |  |  |  |  |  |  |  |  |  |
| Neutral | 1 |  |  | 1 |  |  | 1 |  |  |
| Concordance | 0.9 | 0.9-1.2 | 0.07 | 1.1 | 0.9-1.3 | 0.07 | 1.1 | 0.9-1.3 | 0.1 |
| Discordance | 1.1 | 0.8-1.06 | < **0.05** | 1.2 | 1.03-1.4 | < **0.05** | 1.2 | 1.04-1.4 | < **0.05** |
| *Political ideology* |  |  |  |  |  |  |  |  |  |
| Center | 1 |  |  | 1 |  |  | 1 |  |  |
| Right | 0.8 | 0.7-0.9 | 0.7 | 1.02 | 0.8-1.2 | 0.7 | 1.03 | 0.8-1.2 | 0.6 |
| Left | 1.3 | 1.1-1.5 | < **0.001** | 1.2 | 1.03-1.4 | < **0.05** | 1.2 | 1.05-1.3 | < **0.01** |
| **SOCIO-DEMOGRAPHIC AND HEALTH FACTORS** |  |  |  |  |  |  |  |  |  |
| *Sex* |  |  |  |  |  |  |  |  |  |
| Male | 1 |  |  | 1 |  |  | 1 |  |  |
| Female | 1.6 | 1.4-1.7 | < **0.001** | 1.6 | 1.5-1.8 | < **0.001** | 1.6 | 1.4-1.7 | < **0.001** |
| *Age Groups* |  |  |  |  |  |  |  |  |  |
| Older Adult (60 years old and older) | 1 |  |  | 1 |  |  | 1 |  |  |
| Adult (27 to 59 years old) | 1.7 | 1.4-2.1 | < **0.001** | 1.4 | 1.2-1.7 | < **0.001** | 1.4 | 1.2-1.7 | < **0.001** |
| Young Adult (18 to 26 years old) | 3.5 | 2.9-4.2 | < **0.001** | 3.0 | 2.4-3.8 | < **0.001** | 2.7 | 2.2-3.3 | < **0.001** |
| *Education level* |  |  |  |  |  |  |  |  |  |
| Intermediate | 1 |  |  | 1 |  |  | -- |  |  |
| Basic | 1.09 | 0.8-1.3 | 0.3 | 1.1 | 0.8-1.4 | 0.3 | -- | -- | -- |
| Advanced | 1.05 | 0.9-1.1 | 0.4 | 1.1 | 1.04-1.3 | 0.4 | -- | -- | -- |
| **Employment** |  |  |  |  |  |  |  |  |  |
| Full-time employee | 1 |  |  | 1 |  |  | 1 |  |  |
| Part-time employee | 1.1 | 0.9-1.3 | 0.6 | 0.9 | 0.8-1.1 | 0.8 | 0.9 | 0.8-1.1 | 0.7 |
| Unemployed | 1.7 | 1.5-1.9 | < **0.001** | 1.3 | 1.1-1.4 | < **0.001** | 1.4 | 1.2-1.5 | < **0.001** |
| **Physical activity** |  |  |  |  |  |  |  |  |  |
| Active | 1 |  |  | 1 |  |  | 1 |  |  |
| Inactive | 1.2 | 1.1-1.4 | < **0.001** | 1.5 | 1.1-1.6 | < **0.05** | 1.4 | 1.2-1.6 | < **0.05** |
| **Knowledge about COVID-19 (the ways by which COVID-19 is spread)** |  |  |  |  |  |  |  |  |  |
| Not sure at all | 1 |  |  | 1 |  |  | -- |  |  |
| Not very sure | 0.9 | 0.6-1.4 | 0.5 | 0.8 | 0.5-1.3 | 0.4 | -- | -- | -- |
| Something sure | 0.8 | 0.6-1.2 | 0.1 | 0.7 | 0.5-1.1 | 0.1 | -- | -- | -- |
| Very sure | 0.8 | 0.5-1.1 | 0.2 | 0.7 | 0.5-1.1 | 0.2 | -- | -- | -- |
| **Social capital and family (infected by covid 19)** |  |  |  |  |  |  |  |  |  |
| Not worried at all | 1 |  |  | 1 |  |  | 1 |  |  |
| Not too worried | 1.6 | 0.9-3.05 | 0.2 | 1.04 | 0.6-1.8 | 0.2 | 1.01 | 0.5-1.8 | 0.2 |
| Worry a little bit | 1.2 | 0.7-2.2 | 0.8 | 1.4 | 0.8-2.7 | 0.8 | 1.4 | 0.7-2.6 | 0.9 |
| Very worried | 1.7 | 1.08-2.9 | < **0.05** | 1.5 | 0.9-2.7 | < **0.05** | 1.4 | 0.8-2.6 | < **0.05** |
| **Pandemic quality of life** |  |  |  |  |  |  |  |  |  |
| Excellent | 1 |  |  | 1 |  |  | 1 |  |  |
| Good | 1.9 | 1.5-2.5 | < **0.05** | 2.04 | 1.6-2.6 | < **0.001** | 1.9 | 1.5-2.4 | < **0.001** |
| Bad/Regular | 5.9 | 4.7-7.5 | < **0.001** | 5.6 | 4.4-7.2 | < **0.001** | 5.03 | 4.01-6.3 | < **0.001** |
| **Socio-economic Status** |  |  |  |  |  |  |  |  |  |
| Low | 1 |  |  | 1 |  |  | 1 |  |  |
| Middle | 1.2 | 1.1-1.4 | < **0.05** | 1.3 | 1.1-1.5 | < **0.001** | 1.3 | 1.1-1.5 | < **0.001** |
| High | 1.17 | 1.02-1.3 | < **0.001** | 1.5 | 1.3-1.8 | < **0.001** | 1.6 | 1.4-1.9 | < **0.001** |
| **AIC** |  |  |  |  | 8570.4 |  | 8564.2 |  |  |

OR: Odds Ratio; CI: Confidence Interval.
OR Unadj: Odds Ratio Unadjusted; OR adj: Odds Ratio adjusted

Analysis of sociodemographic and health factors revealed that women [OR: 1.61 (95%CI: 1.46–1.78)], young adults [OR: 2.75 (95%CI: 2.26-3.35)], and inactive participants [OR: 1.40 (95%CI: 0.87–1.16)] were more likely to have alterations in SpMH than men, older adults, and physically active participants, respectively. Moreover, unemployed people [OR: 1.40 (95%CI: 1.24–1.58)], participants who reported bad/regular quality of life during the pandemic [OR: 5.03 (95%CI: 4.01-6.31)], and individuals with high socio-economic status [OR: 1.66 (95%CI: 1.41–1.96)] were more likely to have alterations in SpMH than employed people, people who reported excellent quality of life, and people with low-level socio-economic status, respectively (see Table 3).

As indicated previously, living in Brazil during the pandemic period was associated with the highest likelihood of SpMH alterations compared to living in Mexico. But it is important to look at the effect by country, considering factors like affinity vote for the incumbent and trust in the government. This is because these two factors may be related to the political situation in each country, depending on how its government attempts to mitigate COVID-19.

When considering the interactions between country and vote for the incumbent, on the one hand, and country and trust in government, on the other regression model 3 (see Table 4) revealed a statistically significant association between Brazil and Vote for the incumbent (discordance) and Brazil and Trust in government (no confidence) (p<0.001). As a result, those who lived in Brazil during the pandemic had discordance [OR: 2.05 (95%CI: 1.74– 2.42)] and did not trust their government [OR: 2.10 (95%CI: 1.74–2.42)] were more likely to have SpMH alterations than those who had a neutral position and lived in other Latin American countries.

**Table 4.** Fixed Effects Model including interactions of Country and Vote for the incumbent and Country and Trust in government based on SpMH in Mexican, Colombian, Chilean, and Brazilian adults. January 2021.

| *Variables* | *Model 3 (interactions)* |  |  |
| --- | --- | --- | --- |
|  | *OR adj* | *CI 95%* | *Pr(> \|z\|)* |
| ***Country\*Vote for the incumbent*** |  |  | **< 0.001** |
| Country (Brazil)* Vote for the incumbent (Discordance) | 2.05 | 1.3-3.1 | **< 0.001** |
| Country (Chile)* Vote for the incumbent (Discordance) | 0.9 | 0.5-1.3 | 0.6 |
| Country (Colombia)* Vote for the incumbent (Discordance) | 1.1 | 0.7-1.7 | 0.4 |
| Country (Brazil)* Vote for the incumbent (Concordance) | 1.6 | 1.1-2.4 | **< 0.05** |
| Country (Chile)* Vote for the incumbent (Concordance) | 1.1 | 0.8-1.6 | 0.3 |
| Country (Colombia)* Vote for the incumbent (Concordance) | 1.01 | 0.6-1.4 | 0.8 |
| ***Country\*Trust in government*** |  |  | **< 0.001** |
| Country (Brazil)*Trust (Confidence) | 0.9 | 0.7-1.1 | 0.2 |
| Country (Chile) *Trust (Confidence) | 0.6 | 0.4-0.8 | 0.3 |
| Country (Colombia) *Trust (Confidence) | 0.5 | 0.3-0.6 | 0.3 |
| Country (Brazil) *Trust (No Confidence) | 2.1 | 1.7-2.4 | **< 0.001** |
| Country (Chile) *Trust (No Confidence) | 1.3 | 1.1-1.5 | **< 0.05** |
| Country (Colombia) *Trust (No Confidence) | 1.2 | 0.8-1.7 | **< 0.05** |
| *"\*" this refers to interactions between variables* |  |  |  |
| *Country: Mexico (level reference)* |  |  |  |
| *Vote for the incumbent: Neutral (level reference)* |  |  |  |
| *Trust in government: Neutral (level reference)* |  |  |  |

The probability of SpMH alterations by country was assessed using predictor effects provided by the Effect package of the R software, which produced graphical summaries fitted with linear predictors (see Figure 1), as well as a generalized linear model (GLM) ^19^. We found that, among the four countries, Brazil’s population has the largest likelihood of SpMH alterations among those who were discordant (49.0%) and did not trust their government (44.0%).

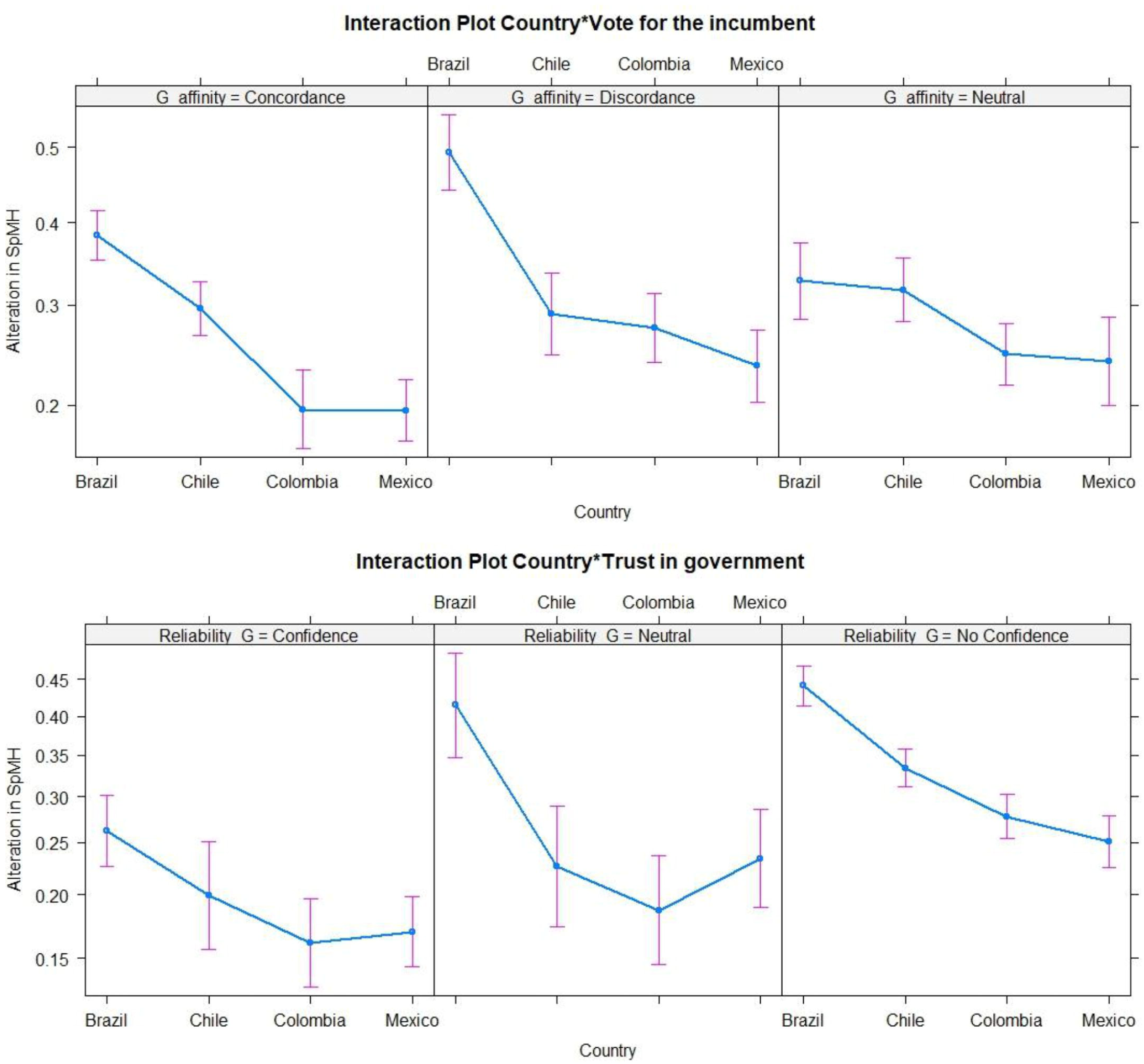

Multivariate model diagnosis showed that the selected model was statistically equivalent to the saturated model (p=0.812), that the model was significant according to the Global Likelihood-Ratio (p<0.01) so that at least one coefficient had a linear relationship with the logit of the outcome, and the Hosmer-Lemeshow test confirmed model fit (p= 0.08). According to the ROC curve, SpMH-affected participants had 72.4% sensitivity and 63.3% specificity.

Finally, 47 data points with a leverage effect were detected from offset residuals bigger than four, and Cook’s distance found no extreme data. We continued using model 2 because the significance of the estimated coefficients did not change (see Equation 2).

*Equation 2. Model 2 (Saturated)*

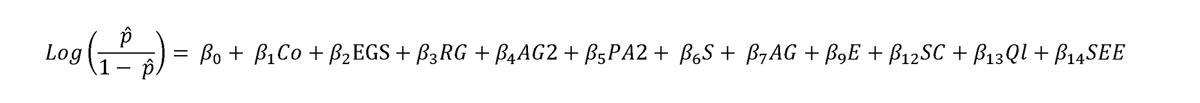

## DISCUSSION

This is the first study in Latin America to examine the prevalence of changes in self-perception of mental health related to the political context of COVID-19 and associated sociodemographic and health factors in Mexico, Colombia, Chile, and Brazil, with the primary objective of obtaining a proportion of 28.8% in these countries.

Multiple research projects investigating politics during pandemics have discovered a notable relationship between health-related occurrences of importance and the implementation of containment measures. Our research in this case found that respondents were more likely to report changes in SpMH if they lacked confidence and disagreed with government policy. According to studies conducted in Latin America, agreement with preventive measures and trust in government were linked to the distribution of social resources during lockdowns; consequently, the number of people with mental health issues increased due to difficulties in obtaining government aid, resulting in increased levels of stress and anxiety among vulnerable populations. The inequities in the allocation of social resources not only affected people’ capacity to comply with suggested preventive measures but also intensified pre-existing discrepancies. The difficulties in obtaining government assistance further highlight the systemic obstacles encountered by communities. The mental health consequences of the pandemic went beyond the acute health crisis, highlighting the importance of inclusive policies and focused treatments to address both the virus and its associated social and mental health impacts in this intricate network of interconnected components. In the wake of lockdowns, it is crucial for society to prioritize resilience and provide fair access to resources in order to effectively respond to future public health issues^6,11^.

The people with a left-wing political orientation were more likely to report having altered SpMH in each of the four countries studied. However, this conclusion for a country like Mexico, where the government was left-wing throughout the COVID-19 outbreak, may be inaccurate. Voting for the incumbent was used to identify countries with a greater likelihood of SpMH alterations in discordant individuals. Some researchers concluded that leftist political identification was statistically associated with psychological stress during the COVID-19 pandemic. Nevertheless, it is crucial to acknowledge the intricate and diverse characteristics of political connections and their influence on mental well-being. Although voting for the incumbent was used as an indicator of SpMH changes, the connection between political ideology and psychological well-being is complex. Further research has emphasized that stress levels during the pandemic were impacted not just by political affiliation, but also by variables such as socioeconomic position, healthcare accessibility, and individual coping strategies. Furthermore, the link between identifying as a lefty politically and experiencing psychological stress highlights the complex relationship between political discussions and mental health results, emphasizing the need for a more thorough investigation into the underlying mechanisms. In the face of global crises and political polarization, it is crucial to take a comprehensive approach to mental health research. This means considering the various factors that influence individuals’ well-being within their political and social environments ^18^. These studies did not suggest that left-leaning and government discordant individuals face greater risks, but they do establish the relationship between vote for the incumbent, political ideology, and mental health in the COVID-19 context.

The third key finding is related to affinity, which is a construction of relationships between personal interests and government policies, so it is a sum of trust and agreeing with government strategies ^5,6^. According to previous analyses of trust in government and vote for the incumbent as well as the fixed effects model, people who had no confidence in their government and lived in Brazil during the COVID-19 pandemic had a higher chance of SpMH alterations than people in other countries in the region.

We identified the concept of social action as a set of policies that governments use to communicate with citizens to create health experiences ^19^. Governments, policymakers, and political leaders have a responsibility to ensure collective and individual health. However, through social action, people became aware that COVID-19 pandemic policies could have negatively affected population health, transcending social unrest and the impact of the alterations in SpMH. Also, some authors considered that the difference between national and local strategies to deal with the COVID pandemic could hurt people’s trust in the government, which could lead to future worries and changes in the mental health of the population. This is known as “punt politics” ^20,21^.

The fourth significant finding of this study is the identification of sociodemographic and health factors associated with an increased risk of SpMH alterations. During the pandemic, the people who suffered the most impact on their mental health were those who had difficulties preserving or finding employment. This outlook about the lack of employability leads to the difficulty of having an income to meet financial needs. After the pandemic progressed, those who were struggling to maintain or obtain job were disproportionately impacted in terms of their mental well-being. The ambiguity regarding job security and financial instability engendered a widespread feeling of apprehension and strain. The connection between challenges in finding employment and mental health issues is complex, since the failure to attain a consistent source of income not only endangers one’s financial security but also affects general contentment and self-worth. The widespread nature of these challenges emphasizes the necessity for comprehensive support systems that tackle both economic and mental health issues ^22^. However, physical activity was identified as a protective variable due to its ability to improve mental health by reducing symptoms like those seen in the study of anxiety, stress, or depression and improving overall emotional well-being. In addition to its physiological advantages, exercise enhances emotional well-being through the stimulation of endorphin release, cultivation of discipline, and enhancement of self-esteem. As societies tackle mental health issues, it is crucial to prioritize regular physical activity, acknowledging the interdependence of physical and mental well-being ^2^.

Finally, the most important findings concern socioeconomic status and quality of life. These are both multidimensional concepts that depend on factors such as life expectancy, income, culture, access to material goods, etc. According to previous research, people in the lowest quintiles of poverty were more likely to develop anxiety and depression because they had a lower quality of life ^23^. Significantly, the countries examined in this study are confronted with significant socioeconomic disparity, which exacerbates the influence of poverty on mental well-being ^22^, which may have altered health-related social gradients and risks. Bad/regular quality of life and high socioeconomic status were more likely to alter their perceptions of their mental health. Our results, different from those in the extant literature, suggest that this association could be explained by high-status people who could not maintain their standard of living due to the pandemic, affecting their quality of life and causing anxiety and depression. Some authors argue that COVID-19 produced an economic contraction that affected entire societies. This advanced viewpoint illuminates the complexity among socioeconomic classes, underlining that the economic difficulties arising from the worldwide health crisis have affected even individuals in historically privileged positions. The notion that COVID-19 triggered a pervasive recession that impacted entire societies is consistent with the broader ramifications of the epidemic on various aspects of life. The economic consequences go beyond individual experiences, infiltrating cultural frameworks and adding to a shared feeling of uncertainty and stress ^24^.

## STRENGTHS AND LIMITATIONS

The participants’ mental health in this study could have been affected before or after the COVID-19’s emergence. This is a limitation of observational study designs due to the fact that they do not track participants over time to estimate the incidence of an outcome as a result of exposure to a specific setting. However, at the same time, it is a strength within a social determinants of health framework, which suggests a multidimensional view of mental health that helps in identifying people at risk based on correlated characteristics ^5^. It is important to note that self-reported data can be affected by reporter bias and overestimation of the true frequencies of health conditions ^25^. This study detected SpMH alterations with a sensitivity of 72.4%, demonstrating that personal perception is an early indicator of mental health changes.

## CONCLUSIONS

Three out of ten survey participants reported sadness, anxiety, or depression during the COVID-19 pandemic. Our study explored how different correlates, especially those of a political nature, exacerbated self-reported mental health outcomes during some of the harshest months of the COVID-19 epidemic in four Latin American countries. Respondents who reported not voting for the incumbent president and those that distrust their government were also more likely to report changes to SpMH. Similarly, implicit risk factors such as unemployment, bad/regular quality of life, or socioeconomic level are also important correlates of changes to SpMH in the four countries under study. In a post-pandemic scenario, our findings could let policymakers create community interventions that include professionals and community mental health actors to reduce SpMH changes.

## STATEMENT ABOUT AUTHORS’ CONTRIBUTIONS

Pablo Roa contributed to the topic, design, data analysis, interpretation, discussion, limitations and strengthening. Guillermo Rosas also contributed to interpretation and discussion. Gloria Isabel Niño Cruz and Sergio Mauricio Moreno López contributed to the data analysis and interpretation. Juliana Mejía Grueso, Haney Aguirre-Loaiza, Javiera Alarcón Aguilar, Rodrigo Reis, Adriano Akira Hino, Fernando López and Deborah Salvo contributed to the interpretation, discussion, limitations, and strengthening. Andrea Ramirez Varela contributed to the topic, design, interpretation, and discussion. All authors reviewed and approved the final version.

## CONFLICT OF INTEREST

There are no conflicts of interest between the authors and the information in this paper.

## Data Availability

All data produced in the present study are available upon reasonable request to the authors

## ACKNOWLEDGMENT

The authors would like to thank the McDonnell International Scholars grant Academy, Universidad de los Andes and Washington University in St. Louis.

## Notes

### Competing Interest Statement

The authors have declared no competing interest.

### Funding Statement

This study did not receive any funding

### Author Declarations

Universidad de los andes, school of medicine, research ethics committee

